# Public adherence to precautionary measures and preventive guidelines against COVID-19 in Sudan: An application of the Health Belief Model

**DOI:** 10.1101/2020.12.25.20248859

**Authors:** Azza Mehanna, Yasir Ahmed Mohammed Elhadi, Don Eliseo Lucero-Prisno

## Abstract

**Background:** Corona virus disease (COVID-19) is highly infectious disease caused by the novel corona virus (SARS-CoV-2). Several public health and social protective measures that may prevent or slow down the transmission of COVID-19 were introduced. However, these measures are unfortunately neglected or deliberately ignored by some individuals.

**Methods:** We did a cross sectional online based survey to identify possible factors influencing intention to adhere to precautionary measures and preventive guidelines against COVID-19 during lockdown periods in Sudan. The questionnaire was used to collect socio-demographic data of study participants, their health beliefs and intention regarding adherence to precautionary measures against COVID-19 based on the constructs of the Health Belief Model.

**Results:** Total of 680 respondents completed and returned the online questionnaire. Significant predictors of intention to adhere to the precautionary measures against COVID-19 were gender (β =3.34, P <0.001), self-efficacy (β= 0.476, P<0.001), perceived benefits (β= 0.349, P<0.001) and perceived severity (β= 0.113, P=0.005). These factors explained 43% of the variance in respondents’ intention to adhere to COVID-19 precautionary measures. Participants who were female, confident in their ability to adhere to the protective measures when available, believing in the benefits of the protective measures against COVID-19 and perceiving that the disease could have serious consequences were more likely to be willing to adhere to the protective measures.

**Conclusion:** Female respondents and respondents having higher self-efficacy, higher perceived benefits and higher perceived severity were more likely to be willing to adhere to the protective measures against COVID-19 in Sudan.

## Introduction

Corona virus disease (COVID-19) is an emerging highly infectious disease caused by the novel corona virus (SARS-CoV-2)[1]. The virus was first detected in Wuhan, China[2] on December 2019. In the last two decades Corona viruses have caused two large-scale pandemics; SARS[3] and MERS[4].Several public health and social measures that may prevent or slow down the transmission of COVID-19 were introduced; including case isolation, identification and follow-up of contacts, adequate environmental disinfection, and use of personal protective equipment (PPE)[5].By 27 August, 2020 the virus has reached 214 countries resulted in 24.2 million cases and 826000 confirmed deaths.

At that early stage of virus spread, there was no approved vaccine preventing corona virus disease. The best prevention was to avoid exposure to the virus[6]. In response to this Pandemic, Almost all countries imposed lockdown policy to slow the spread of the virus and introduced measures that may reduce the risk of exposure, these include: use of face masks; regular hand washing with soap or disinfection with hand sanitizer; avoidance of contact with infected people and complying with social distancing measures in crowded places; and avoid touching eyes, nose, and mouth with unwashed hands [7, 8]. However, these guidelines, which were introduced to slow the spread of COVID-19 and contribute to public well-being, are unfortunately neglected or deliberately ignored by some individuals [9].

Health Belief Model [10] was originally formulated to explain different preventive health behaviors. According to this model, the individual’s health behavior depends on an individual’s perception of being at risk to get the disease, perceived severity of the disease, perceived benefits and perceived costs of taking a particular health action [11] and feeling capable of implementing the desired behavior to achieve results[12]. This study aims to identify the possible factors influencing Sudanese people’s intention to adhere protective measures and preventive guidelines (staying at home, wearing masks/gloves and following social distancing measures) using domains (constructs) of the Health Belief Model.

## Methods

### Study Design and Sampling

A cross-sectional design was used to study participants’ intention to adhere to protective measures against COVID-19. The study was conducted in Sudan in 2020.The study was conducted on 625 participants based on the assumption that intention to adhere to protective measures =50%, precision =5% and alpha = 0.05. Convenience internet-based sampling technique was used.

### Data Collection

Using Google forms, an online structured questionnaire was developed by the researchers based on review of previous literature. The questionnaire link was distributed to the participants through social media such as: Email, Facebook, WhatsApp, Twitter .Etc. The questionnaire was used to collect socio-demographic data of study participants, their health beliefs and intention regarding adherence to precautionary measures against COVID-19. The precautionary measures and preventive guidelines addressed in this study were: staying at home during lockdown periods, wearing masks/gloves and practicing social distancing on going out.

### Constructs of the Health Belief Model (HBM)

the scale measuring HBM constructs consists of 5 subscales measuring perceived susceptibility & severity of COVID-19, participant’s perceived benefits and barriers to adhere to precautionary measures and self-efficacy in adhering to preventive guidelines. Responses to scale items were scored on a five-point Likert scale ranging from 0 (strongly disagree) to 4 (strongly agree). The score was reversed for some items, it was calculated for each subscale, converted to percentage and categorized into high (> 66.67%), moderate (33.33% to 66.67%) and low (< or equal to 33.33%).

### Intention of respondents to Adhere to Precautionary Measures and Preventive Guidelines

It was assessed using two statements: “In the coming period, I will try to stay at home and not go out unless necessary”, and “In the coming period, I will try to adhere to the protective measures (wearing a mask, gloves and social distancing) when going out”. Responses to scale items were scored on a five-point Likert scale ranging from 0 (strongly disagree) to 4 (strongly agree). The score was calculated and converted to percentage and categorized into high (> 66.67%), moderate (33.33% to 66.67%) and low (< or equal to 33.33%).

The internal consistency of HBM subscales and the scale for intention to adhere to precautionary measure were determined by Cronbach’s alpha coefficient. Cronbach’s alpha coefficients were 0.862 (perceived susceptibility),0.798 (perceived severity), 0.799 (perceived benefits), 0.793 (perceived barriers), 0.757(self-efficacy), 0.798 (perceived severity) and 0.823 (Intention to adhere to precautionary measures).

### Statistical analysis of the data

Data were fed to the computer and analyzed using IBM SPSS software package version 20.0(Armonk, NY: IBM Corp). Qualitative data were described using number and percent. Quantitative data were described using mean and standard deviation. Significance of the obtained results was judged at the 5% level. Pearson’s coefficient was used to correlate between the different variables used in the study. Linear regression analysis was performed to detect the significant predictors of intention to adhere to precautionary measures.

### Ethical consideration

This study was approved by the Ethics Committee of the High Institute of Public Health, Alexandria University, Egypt in 2020. The questionnaire was preceded by a cover letter explaining the aim of the study followed by an invitation to participate. The anonymity and confidentiality of participants were guaranteed. Online submission of the questionnaire was considered as consent to participate in the study.

## Results

### Socio-demographic characteristics of the respondents

A Total of 680 Sudanese has responded to the questionnaire. The mean age of participants was 26.7 (±7.95 years), 56.8% of participants were females, 83.7% were living in Khartoum (the capital of Sudan), 79% were single and77.6% were university undergraduates. The monthly income of about half the participants (49.6%) was just enough (Table 1).

**Table (1):** Socio-demographic characteristics of study participants.

| Socio-demographic characteristics | Number | % |
| --- | --- | --- |
| **Age** |  |  |
| <20 | 58 | 8.5 |
| 20 - <40 | 566 | 83.2 |
| 40 - <60 | 52 | 7.6 |
| 60+ | 4 | 0.6 |
| Mean ± SD | 26.70±7.95 | |
| **Sex** |  |  |
| Male | 294 | 43.2 |
| Female | 386 | 56.8 |
| **Residence** |  |  |
| Northern State | 16 | 2.4 |
| Red Sea State | 32 | 4.7 |
| Al-Gezira State | 51 | 7.5 |
| Khartoum State | 569 | 83.7 |
| River Nile State | 12 | 1.8 |
| **Marital status** |  |  |
| Single | 537 | 79.0 |
| Married | 131 | 19.3 |
| Divorced/ Widowed | 12 | 1.7 |
| **Education** |  |  |
| High school | 35 | 5.1 |
| University undergraduate | 528 | 77.6 |
| University graduate | 117 | 17.2 |
| **Income** |  |  |
| Not enough | 104 | 15.3 |
| Just enough | 337 | 49.6 |
| Enough and saving | 239 | 35.1 |

### Mean percent scores of the Health Belief model domains and Respondent’s intention to adhere to precautionary measures

The mean percent scores (MPS) of all the domains of the HBM –except for perceived barriers were pertinently high indicatingthat participants strongly believedin being suceptible to COVID-19 (80.48%) and that the disease could have severe consequences(84.99%). Participants also perceived that adherence to the protective measures such as social distancing and wearing masks was important and beneficial (93.65%)and were confident in their ability to adhere to these protective measures(81.01%) if they were available. Nevertheless, the MPS of perceived barriers was moderately high (61.48%) denoting that several barriers to the adherence to protective measures were reported by a significant proportion of participants.The MPS of intention to adhere with authority guidlines was high (87.21%) reflecting participants’ strong will to adhere to the protective measures against COVID-19 (Figure 1).

**Figure 1:**
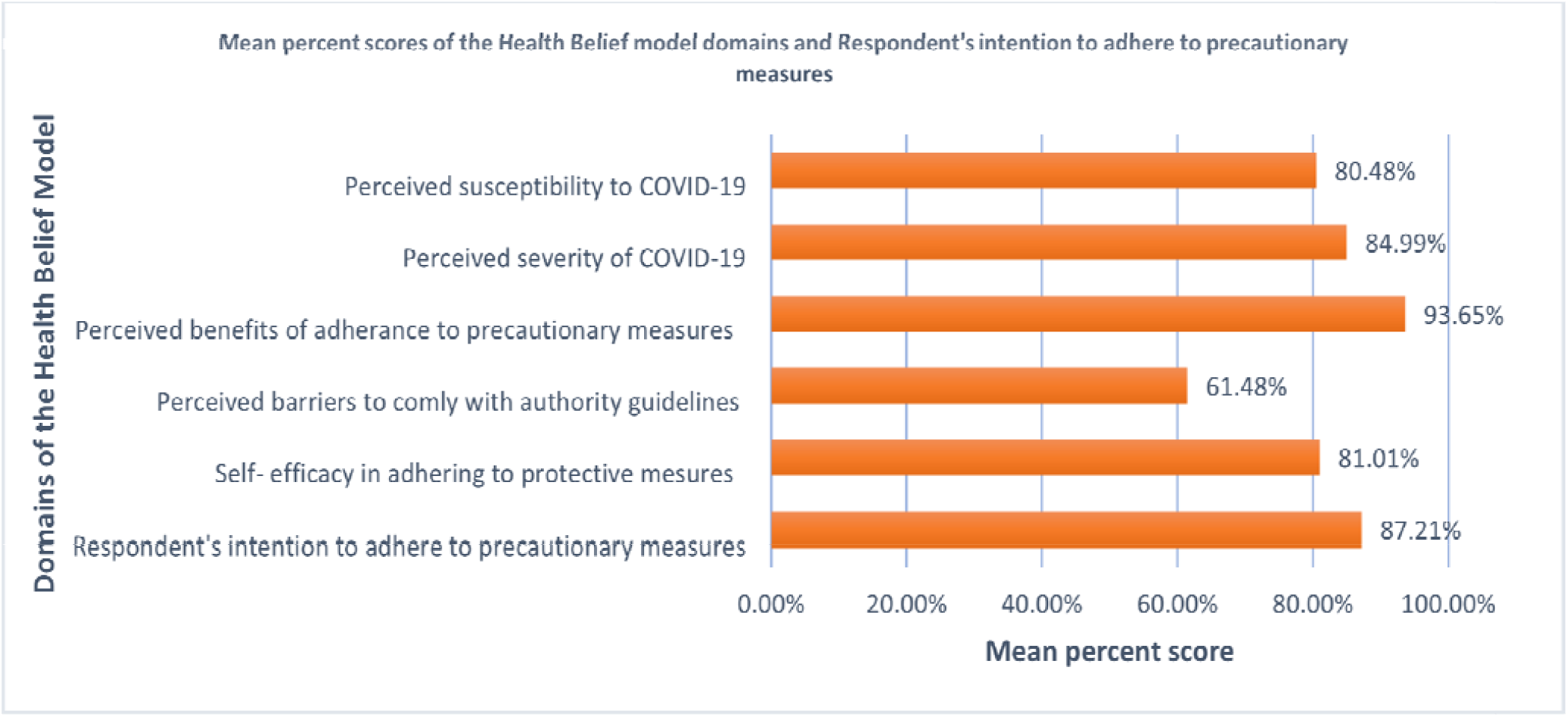
Mean percent scores of the Health Belief model domains and Respondent’s intention to adhere to precautionary measures.

### Perceived susceptibility and perceived severity (perceived threat) of respondents to COVID-19

Most of the participants agreed/strongly agreed on being at risk of getting COVID-19 (71.3%) and on the possibility of getting infected when being in contact with a person not showing the symptoms of the disease (87.6%) meanwhile, they disagreed/strongly disagreed on the negative statement “Covid-19 disease is not transmitted through contact with surfaces or tools”(80.4%).However, more than one third (35.6%) of participants were not sure whetherthe hot climate in Sudan killed the virus and thus reduced the possibility of infection. More than 90% of the participants agreed/strongly agreed that COVID-19 is spreading rapidly and that it may lead to dealth. More than 90% agreed/srongly agreed that adherence to protective measures reduced the number of cases and decreased spread of the disease) (Table 2).

**(Table 2):**
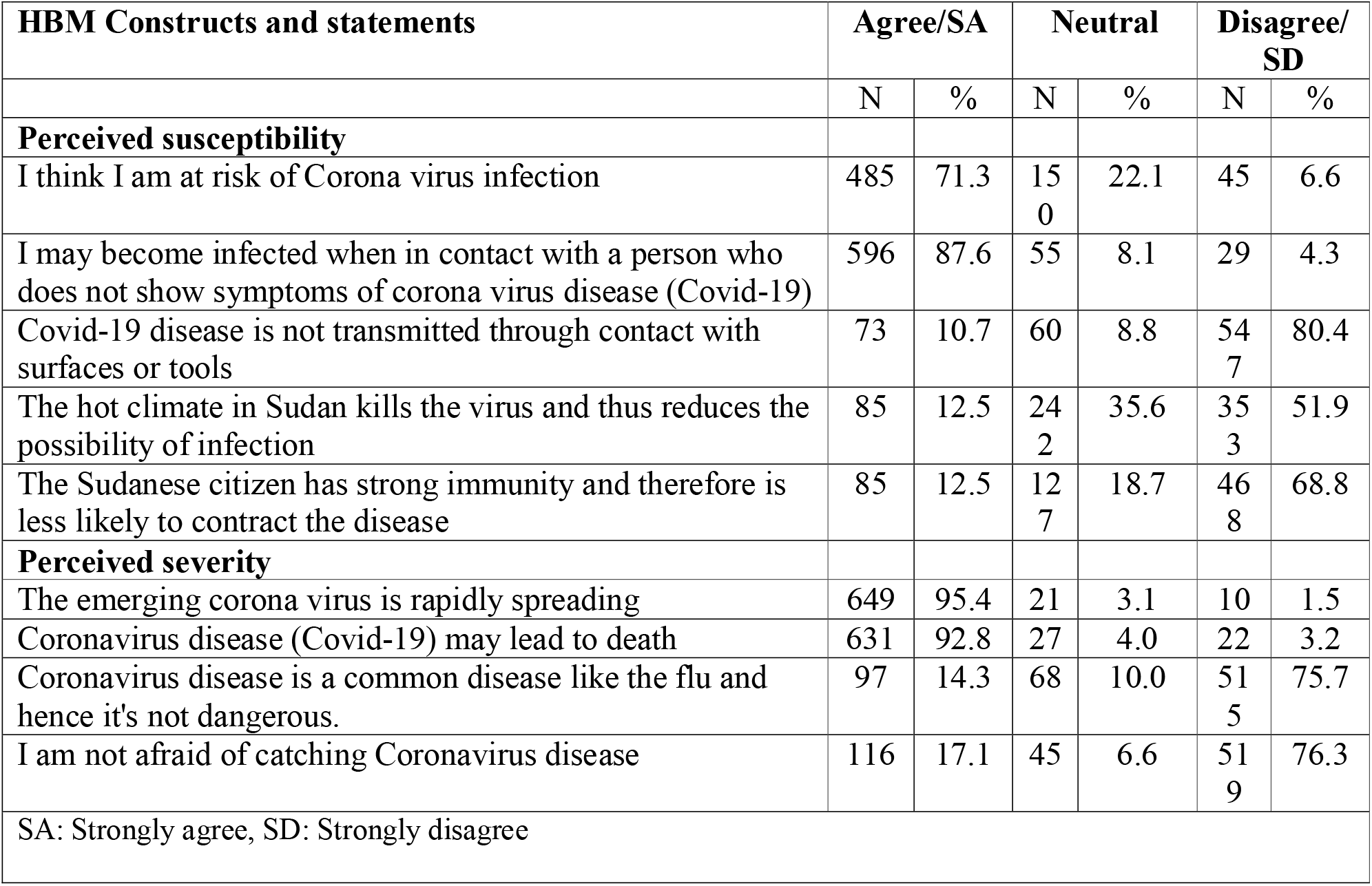
Perceived susceptibility and perceived severity (perceived threat) of respondents to COVID-19.

| HBM Constructs and statements | Agree/SA |  | Neutral |  | Disagree/SD |  |
| --- | --- | --- | --- | --- | --- | --- |
|  | N | % | N | % | N | % |
| **Perceived susceptibility** |  |  |  |  |  |  |
| I think I am at risk of Corona virus infection | 485 | 71.3 | 150 | 22.1 | 45 | 6.6 |
| I may become infected when in contact with a person who does not show symptoms of corona virus disease (Covid-19) | 596 | 87.6 | 55 | 8.1 | 29 | 4.3 |
| Covid-19 disease is not transmitted through contact with surfaces or tools | 73 | 10.7 | 60 | 8.8 | 547 | 80.4 |
| The hot climate in Sudan kills the virus and thus reduces the possibility of infection | 85 | 12.5 | 242 | 35.6 | 353 | 51.9 |
| The Sudanese citizen has strong immunity and therefore is less likely to contract the disease | 85 | 12.5 | 127 | 18.7 | 468 | 68.8 |
| **Perceived severity** |  |  |  |  |  |  |
| The emerging corona virus is rapidly spreading | 649 | 95.4 | 21 | 3.1 | 10 | 1.5 |
| Coronavirus disease (Covid-19) may lead to death | 631 | 92.8 | 27 | 4.0 | 22 | 3.2 |
| Coronavirus disease is a common disease like the flu and hence it's not dangerous. | 97 | 14.3 | 68 | 10.0 | 515 | 75.7 |
| I am not afraid of catching Coronavirus disease | 116 | 17.1 | 45 | 6.6 | 519 | 76.3 |
| SA: Strongly agree, SD: Strongly disagree |  |  |  |  |  |  |

### Respondent’s Self-efficacy, perceived barrier and perceived benefits to adhere to precautionary measures and preventive guidelines against COVID-19

A great majority of the participants believed that they were capable of staying at home for the entire period of time specified by the ministry of health, and only go out if necessary (78.7%) and were able to commit to wearing the mask (if available) whenever they went out (91.6%). Regarding perceieved barriers, bring the needs of the family was the most frequently reported factor hindering participants from satying at home (85.1%) followed by work requirements (62.4%) then feeling bored (46.8%), while disagreements between family members at home was the least mentioned barrier to staying at home (17.6%). Concerning factors hindering adherence to protective measures outside home,the absenceof rules governing the distance between people in crowded places such as markets and means of public transportation was the barrier most agreed/strongly agreed upon(91.8%) followed by the unavailability and high prices of gloves/masks (76.3% and 72.6% respectively) (Table 3).

**(Table 3):**
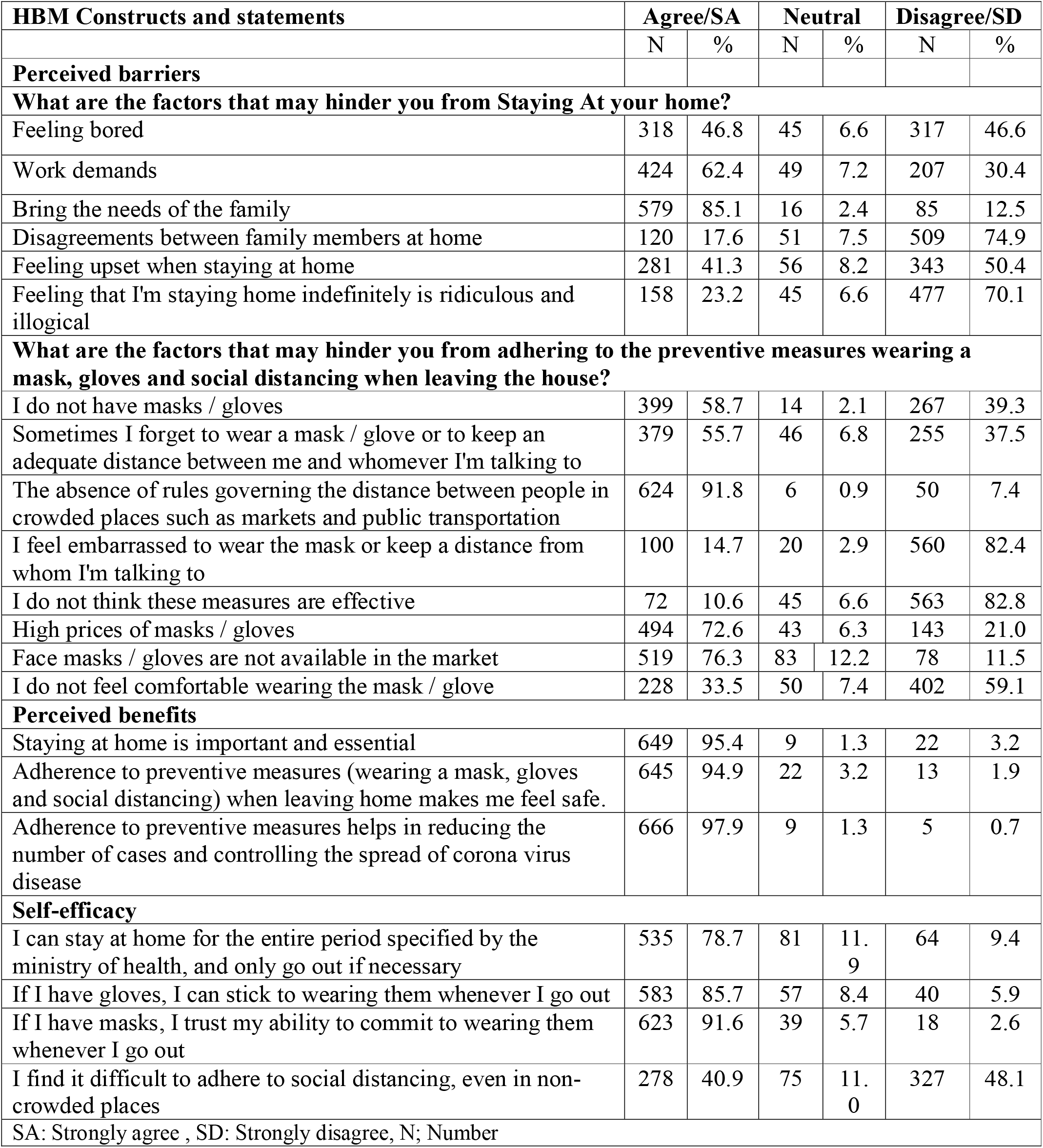
Respondent’s Self-efficacy, perceived barrier and benefits of adherence to precautionary measures and preventive guidelines against COVID-19.

### Correlation between constructs of the Health Belief Model and intention of respondents to adhere to precautionary measures

Intention to adhere to preventive guidelines was significantly correlated with all the parameters of HBM model. The five parameters of the model were significantly positively correlated except for perceived barriers which, expectedly, showed a significant negative correlation with intention and with all the other parameters of the HBM. Self-efficacy had the strongest positive correlation with intention to adhere to preventive measures (r =0.589) followed by perceived benefits (r =0.490). Intuitively, this means that the more confident the person is in his ability to perform the behavior (adhere to protective measures), the more likely he feels he would perform the behavior. However, intention to adhere to protective measures is influenced by other factors as well including perceived benefits and perceived severity (Table 4).

**(Table 4):**
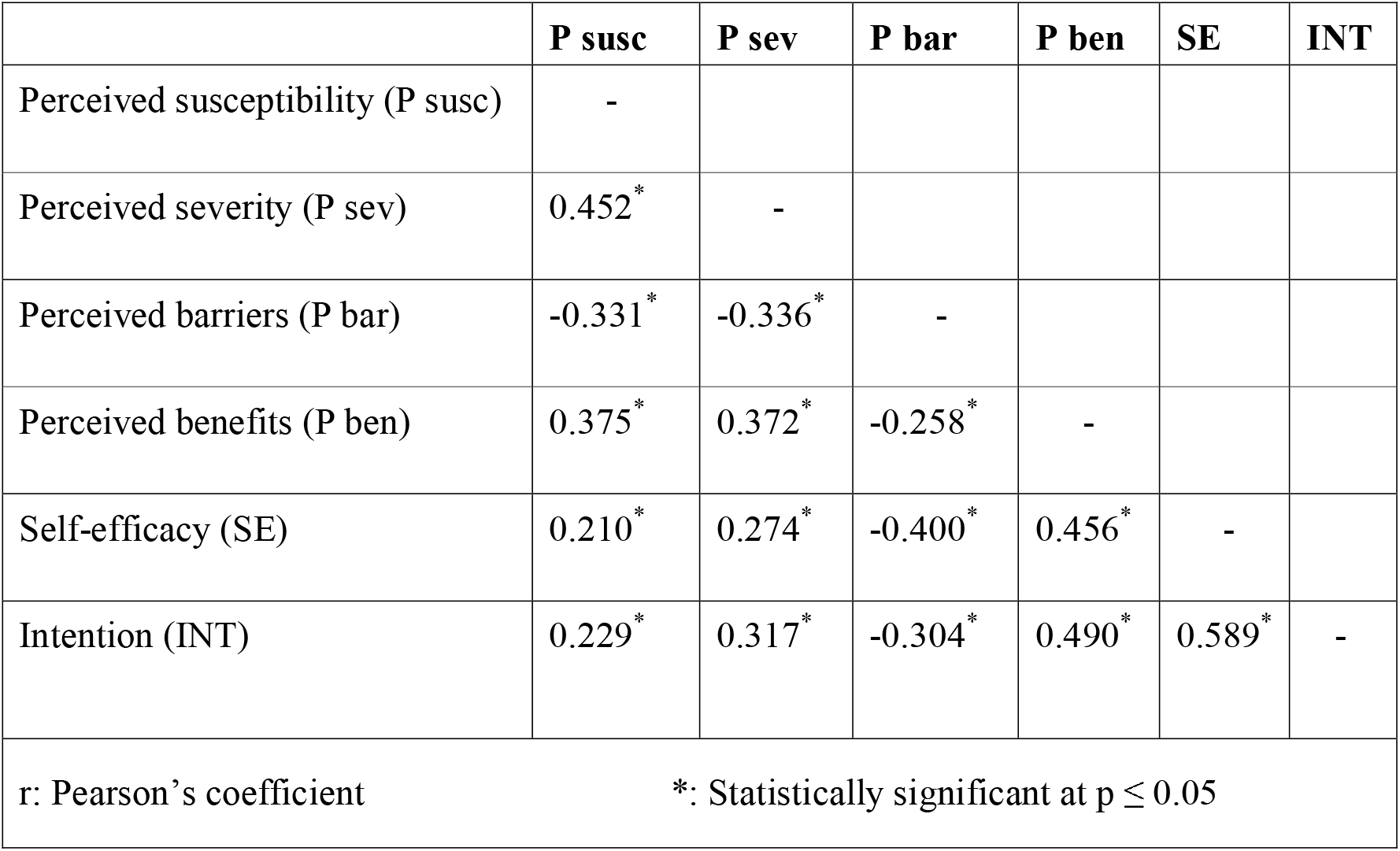
Correlation between constructs of the Health Belief Model and intention of respondents to adhere to precautionary measures.

### Predictors of participant’s intention to adhere to precautionary measures

Table 5 shows the multivariate linear regression analysis of respondent’s intention to adhere to the protective measures, with some independent variables. After adjustment for all possible confounders, it was found that the significant predictors of intention were gender (β =3.34, P<0.001), self-efficacy (β= 0.476, P<0.001), perceived benefits (β= 0.349, P<0.001) and perceived severity (β= 0.113, P=0.005). These factors explained 43% of the variance in participants’ intention to adhere to the protective measures. Participants who were female, confident in their ability to adhere to the protective measures when available, believing in the benefits of the protective measures against COVID-19 and perceiving that the disease could have serious consequences were more likely to be willing to adhere to the protective measures (Table 5).

**(Table 5):** Summary of multivariate linear regression analysis for the parameters affecting respondent’s intention to adhere to precautionary measures.

| Parameters | #Multivariate |  |
| --- | --- | --- |
|  | p | B (95% C. I) |
| Sex (1=male/ 2= female) | <0.001<sup>*</sup> | 3.342 (1.592 – 5.091) |
| Income | 0.196 | 0.858 (-0.443 – 2.159) |
| Perceived susceptibility | 0.489 | 0.027 (-0.104 – 0.050) |
| Perceived severity | 0.005<sup>*</sup> | 0.113 (0.035 – 0.192) |
| Perceived barriers | 0.730 | -0.013 (-0.087 – 0.061) |
| Perceived benefits | <0.001<sup>*</sup> | 0.349 (0.250 – 0.449) |
| Self-efficacy | <0.001<sup>*</sup> | 0.476 (0.400 – 0.552) |
| R<sup>2</sup>=0.434, F=73.522<sup>*</sup>, p<0.001<sup>*</sup> |  |  |

## Discussion

This study focused on identifying the health beliefs influencing intention to adhere to precautionary behavior against the novel corona virus pandemic in Sudan. While suffering the burdens of civil war, political instability and economic meltdown [13]. Sudan, like all other countries, was hit by the COVID-19 pandemic. Despite the unfavorable political and economic conditions and to our surprise respondents showed high intention to adhere to the protective measures against COVID-19. Most of the respondents were relatively young which offers some explanation to their readiness to accept the change and fight their battle against COVID-19. Evidently, females were more likely to be willing to adhere to the protective measures against the disease. This finding was supported by several studies [14-18] which all agreed that females were more compliant to health-related guidelines than males. Intuitively, health educators should invest more effort in educating females who would, in turn, act as change agents influencing their social networks.

Regarding health beliefs, participants scored high on their health beliefs about adherence to the protective measures. It seems that the worldwide awareness-raising campaigns played an important role in increasing peoples’ knowledge of the virus and helped them acquire a more negative attitude towards the disease and a more positive attitude towards adopting the protective measures against the disease. Hence, most participants in the present study believed in being susceptible to the disease whether directly through their contact with infected people or indirectly through their contact with virus-contaminated surfaces or tools. However, a non-negligible proportion of participants neither agreed nor disagreed that the hot climate in Sudan killed the virus and believed that the Sudanese citizen had strong immunity and therefore were less likely to contract the disease, without documented scientific evidence. Such findings which have been previously reported [13] should draw the attention of health authorities in Sudan to the need of providing credible information to correct the misunderstanding and clear the ambiguity people might have about the disease. Nonetheless, it seems that the role of perceived susceptibility was not very evident in enhancing participants’ intention to adhere to the protective measures.

An intriguing result was that among health beliefs, self-efficacy-and not perceived barriers-was the most significant predictor of intention. Feeling capable of adhering to protective measures against COVID-19 has strongly influenced participants’ intention to do so. This finding highlights the importance of enhancing people’s confidence in their ability to adhere to protective measures by disseminating credible information about these measures: what they are, where to be found, when and how to be used. Moreover, they should be affordable and accessible. Additionally, enforcing rules concerning social distancing and wearing masks would transform these precautionary measures into accepted social norms, thus, encouraging people and giving them the confidence to adhere to such measures. In his Social Cognitive theory, Albert Bandura described self-efficacy as the most important pre-requisite for behavior change[19, 20]. Indeed, a considerable amount of research revealed that self-efficacy was a significant predictor of health behavior [21-23] lending support to our findings.

Examination of correlation analyses revealed that intention to adhere to the protective measures was also influenced by perceived benefits of the protective measures and perceived severity of the disease, in the mentioned order. These findings should direct our efforts to the priority areas needed to be addressed while educating people about the importance of adherence to the protective measures. Undoubtedly, knowing the role of each protective measure in protecting people from being infected by the novel Corona virus and from the possible serious consequences of the disease would encourage them to adhere to these protective measures. Previous research has shown perceived benefits and perceived severity to be significant predictors of preventive health behavior [24 -27].

Several barriers to adherence were documented in the current study, most important of which were the high prices of masks/gloves, their unavailability in the market and the absence of rules governing the distance between citizens in public places and public transportation. As regards staying at home, the most important constraints were bringing family needs and work demands. Despite not being a significant predictor of intention, these barriers should not be overlooked. In fact, resolving these barriers would be an avenue to enhancing people’s self-efficacy and perceived controllability of adherence behavior. Ultimately, emphasis on some HBM constructs does not mean that we should ignore other constructs to achieve synergetic effect. In contradiction to our findings, several studies found perceived barriers to be a significant predictor of preventive health behaviors including immunization and screening behaviors [28 -30].

### Implications for research and practice

This research could be an important guide in designing health education messages to enhance adherence to the protective measures againstCOVID-19. Educational messages should essentially focus on improving people’s self-efficacy in adhering to the protective measures. Moreover, more efforts should be invested in targeting females who would act as influential change agents among their social networks.

### Limitations

Cues to action were not assessed in this study to avoid a long questionnaire, however, knowing the triggers instigating participants to adhere to the protective measures would have certainly added to this work. We used the convenience sampling technique; hence generalization of data would not be possible. Nonetheless, the present research was able to shed important light on some of the factors influencing participants’ intention to adhere to the protective measures in Sudan. A subjective tool -self-report questionnaire -was used to collect data; therefore, participants’ responses might be affected by social desirability

## Conclusion

Despite the unfavorable political and economic conditions in Sudan our respondents showed high intention to adhere to the protective measures and health authority guidelines against COVID-19. female respondents and respondents having higher self-efficacy, higher perceived benefits and higher perceived severity were more likely to be willing to adhere to the protective measures against COVID-19 in Sudan. The Health Belief model was a useful tool in predicting the beliefs affecting respondent’s intention to adhere to the protective measures.

### What is known about this topic

- COVID-19 pandemic has posed unprecedented challenges and impact on all aspects of life.
- Protective measures and precautionary guidelines against COVID-19 are being neglected by some individuals in the community.
- The Health Belief Model has been applied widely to explain different preventive health behaviors.

### What this study adds

- Going to bring the needs of the family was the most frequently reported factor hindering respondents from satying at home during lockdown periods in sudan followed by work requirements and feeling bored.
- Absence of rules governing the distance between people in crowded places such as markets and means of public transportation, unavailability and high prices of gloves/masks were the most barriers hindering respondents froms adhering to precautionary measures outside home.
- Based on our findings, health education messages regarding COVID-19, should essentially focus on improving people’s self-efficacy in adhering to protective measures and precautionary guidelines against COVID-19.

## Data Availability

Data and questionnaire are available from the corresponding author upon request.

## Authors’ contributions

The concept for this research was developed by YE and AM. YE and AM designed the Data collection tool, collected the data, developed the draft and prepared the manuscript with an important contribution from DL. All the authors have read and agreed to the final manuscripts.

## Acknowledgements

None.

